# Expression of specific HLA class II alleles is associated with an increased risk for active tuberculosis and a distinct gene expression profile

**DOI:** 10.1101/2022.07.06.22277293

**Authors:** Leila Y. Chihab, Rebecca Kuan, Elizabeth J. Phillips, Simon A. Mallal, Virginie Rozot, Mark M. Davis, Thomas J. Scriba, Alessandro Sette, Bjoern Peters, Cecilia S. Lindestam Arlehamn, the SATVI Study Group

**Author notes:** Corresponding author Cecilia S. Lindestam Arlehamn.

## Abstract

Several HLA allelic variants have been associated with protection from or susceptibility to infectious and autoimmune diseases. Here, we examined whether specific HLA alleles would be associated with different *Mycobacterium tuberculosis (Mtb)* infection outcomes. The HLA alleles present at the -A, -B, -C, -DPA1, -DPB1, -DQA1, -DQB1, -DRB1, and -DRB3/4/5 loci were determined in a cohort of 636 individuals with known *Mtb* infection outcomes from South Africa and the US. Among these individuals, 233 were QuantiFERON (QFT) negative, and 433 were QFT positive, indicating *Mtb* exposure. Of these, 99 QFT positive participants either had active tuberculosis (TB) upon enrollment or were diagnosed in the past. We found that DQA1*03:01, DPB1*04:02, and DRB4*01:01 were significantly more frequent in individuals with active TB (susceptibility alleles), as judged by Odds Ratios and associated p-values, while DPB1*105:01 was associated with protection from active TB. Peripheral blood mononuclear cells (PMBCs) from a subset of individuals were stimulated with *Mtb* antigens, revealing individuals who express any of the three susceptibility alleles were associated with lower magnitude of responses. Furthermore, we defined a gene signature associated with individuals expressing the susceptibility alleles that was characterized by lower expression of APC-related genes. In summary, we have identified specific HLA alleles associated with susceptibility to active TB and found that the expression of these alleles was associated with a decreased *Mtb*-specific T cell response and a specific gene expression signature. These results will help understand individual risk factors in progressing to active TB.

## Introduction

*Mycobacterium tuberculosis* (*Mtb*), the cause of tuberculosis (TB) disease, infects about one-quarter of the world’s population and causes approximately 1.5 million deaths annually ^1^. Although *Mtb* infection is fairly prevalent worldwide, less than 10% of infections lead to active tuberculosis (ATB) disease^1,2^. In some cases individuals with *Mtb* infection can completely clear all living bacteria, while in most *Mtb* infection cases the bacteria persist and these individuals have an increased risk of bacterial reactivation leading to ATB ^3^. Latent infection can last weeks, months, or years with the likelihood of progression to ATB being highest within the first two years following infection. Certain conditions can increase the likelihood of progression to ATB disease (e.g. HIV co-infection, immune suppression therapy, malnutrition, and diabetes) ^4^. To reduce the incidence of TB the WHO has identified a strategy which includes identification and treatment of individuals with ATB, screening of individuals at high risk, as well as preventative therapy to those at high risk of progression to ATB ^5^. Importantly, the biological correlates that distinguish between different *Mtb* infection outcomes have yet to be fully elucidated and thus there is a need for improved prognostic and diagnostic tests to identify those at risk for ATB.

Human Leukocyte Antigens (HLAs) are heterodimeric proteins that play a critical role in the immune recognition of antigens. HLA molecules present pathogen-derived peptides (epitopes) for recognition by T cells ^6^. HLA molecules are grouped into class I vs. class II, each encompassing several highly polymorphic loci with more than 15,000 different possible HLA allelic variants ^7^. In general, different HLA allelic variants present distinct epitope repertoires, and this high level of diversity results in a highly diverse set of peptide epitopes being presented in different individuals in a given population. In this context, genetic analyses and genome wide association studies (GWAS) have demonstrated that the presence of specific HLA allelic variants can exert either a protective or susceptibility effect for different autoimmune or infectious diseases ^8-14^.

Here, we examined whether any specific HLA alleles are associated with ATB and thus could be used to identify individuals at risk for ATB, and whether expression of these HLA alleles can be linked to differences in the *Mtb*-specific immune response.

## Materials and Methods

### Ethics Statement

This research was performed in accordance with approval from the Human Research Ethics Committees of the University of Cape Town (Cape Town, South Africa) and the La Jolla Institute for Immunology (La Jolla, USA). All participants provided written informed consent before participation. Adolescents provided written informed assent, with a parent or legal guardian providing written informed consent.

### Study Participants

Study participants (n=620) from the Worcester region of South Africa were investigated. Participants were divided into a healthy cohort of QFT positive (n=318) and negative (203) individuals (QuantiFEROn-TB Gold In-Tube, Cellestis) and into individuals with ongoing (sputum XpertMTB/RIF-positive (Cepheid, Sunnyvale, CA)) TB disease (n=28) or a self-reported history of individuals with previously diagnosed active TB who underwent TB treatment (n=71). We also enrolled QFT+ participants (n=16) at the University of California, San Diego Anti-Viral Research Center (San Diego, USA). QFT status and whether individuals have or have had active TB is listed in **Supplementary table 1**.

### PBMC Isolation

PBMC were purified from whole blood either using CPT tubes (BD), for the adolescents, or layered onto Ficoll (for adults) using 50ml Leukosep tubes (Greiner) by density-gradient centrifugation on Ficoll, according to the manufacturer’s instructions. Cells were suspended in FBS containing 10% (vol/vol) DMSO and cryopreserved in liquid nitrogen. The cryopreserved cells from South Africa were shipped to LJI for analysis.

Cryopreserved PBMC were quickly thawed by incubating each cryovial at 37°C for 2 min, and cells transferred to cold medium (RPMI 1640 with L-glutamin and 25mM HEPES; Omega Scientific), supplemented with 5% human AB serum (GemCell), 1% penicillin streptomycin (Life Technologies), 1% glutamax (Life Technologies) and 20 U/ml benzonase nuclease (MilliporeSigma). Cells were centrifuged and resuspended in medium to determine cell concentration and viability using trypan blue and a hematocytometer.

### Peptides and other stimuli

Peptides were synthesized as crude material on a small (1 mg) scale by A&A LLC (San Diego). Peptides were pooled into peptide pools, re-lyophilized and reconstituted at a concentration of 0.7mg/ml for MTB300 (PMID27409590). *Mtb* whole cell lysate (WCL) from strain H37Rv was obtained from BEI Resources, NIAID, NIH (NR-14822).

### HLA typing

Participants were HLA-typed at the La Jolla Institute, by the American Society for Histocompatibility and Immunogenetics (ASHI)-accredited laboratory at Murdoch University (Western Australia), or Immucor, Sirona Genomics, CA.

Typing at LJI was performed by next-generation sequencing ^15^. Specifically, amplicons were generated from the appropriate class II locus for exons 2 through 4 by PCR amplification. From these amplicons, sequencing libraries were generated (Illumina Nextera XT) and sequenced with MiSeq Reagent Kit v3 as per the manufacturer’s instructions (Illumina). Sequence reads were matched to HLA alleles and participant genotypes were assigned.

Typing in Australia for class I (HLA A, B, C) and class II (DQA1, DQB1, DRB1, 3, 4, 5, DPB1) was performed using locus-specific PCR amplification of genomic DNA. Patient-specific, barcoded primers were used for amplification. Amplified products were quantitated and pooled by participant and up to 48 participants were pooled. An indexed (8 indexed MiSeq runs) library was then quantitated using Kappa universal QPCR library quantification kits. Sequencing was performed using an Illumina MiSeq using 2×300 paired-end chemistry. Reads were quality-filtered and passed through a proprietary allele calling algorithm and analysis pipeline using the latest IMGT HLA allele database as a reference. The algorithm was developed by E.J.P. and S.A.M. and relies on periodically updated versions of the freely available international immunogenetics information system (http://www.imgt.org) and an ASHI-accredited HLA allele caller software pipeline, IIID HLA analysis suite (http://www.iiid.com.au/laboratory-testing/). Typing at Immucor, Sirona Genomics was performed using the MIA FORA NGS kit and analysis software (Immucor, Inc.) as previously described ^16^.

The HLA type of each subject is listed in **Supplementary table 1**.

Population coverage of HLA alleles were calculated using the population coverage tool from the Immune epitope database (http://www.iedb.org).

### ELISPOT assay

PBMCs were thawed and antigen-specific cellular responses were measured by IFNγ ELISPOT assay with all antibodies and reagents from Mabtech (Nacka Strand, Sweden). Plates (96-well Immobilion-P; Millipore) were coated overnight at 4°C with 5µg/ml anti-IFNI (1-D1K; Mabtech). Briefly, 200,000 cells were plated in each well of the pre-coated plate and were stimulated in triplicate with 10µg/ml *Mtb* whole cell lysate, 1 µg/ml MTB300, 10µg/ml PHA as a positive control, or DMSO corresponding to the concentration present in the peptide pool. After 20h incubation at 37°C, wells were washed with PBS/0.05% Tween 20 and incubated with biotinylated anti-IFN□ (7-B6-1; Mabtech) for 2h. Spots were developed using Vectastain APC peroxidase (Vector Laboratories) and 3-amino-9-ethylcarbazole (Sigma-Aldrich). Spots were counted by computer-assisted image analysis (AID iSpot; Aid Diagnostica). Responses were considered positive if the net spot-forming cells (SFC) per 10^6^ PBMC were ≥20, the stimulation index ≥2, and p≤0.05 (Student’s t-test, mean of triplicate values of the response against relevant stimuli vs. the DMSO control).

### Cell sorting and RNA purification

PBMCs were thawed and added at a density of 1×10^6^ cells per well to a round-bottom 96-well plate. They were stimulated for 24 hrs at 37°C with MTB300, *Mtb* whole cell lysate, anti-CD28 (1µg/ml, CD28.2; eBioscience) together with anti-CD3 (1µg/ml pre-coated overnight at 4°C, UCHT1; BioLegend), as well as left unstimulated as a negative control. After 24 hrs, cells were resuspended in PBS with 10% (v/v) FBS and incubated at 4°C for 10 min. Cells were then stained with fixable viability dye eFluor506 (eBioscience) and an antibody mixture containing anti-human CD3-AF700 (UCHT1; ThermoFisher), CD4-APCeFluor780 (RPA-T4; eBioscience), CD8-BV650 (RPA-T8; BioLegend), CCR7-PerCPCy5.5 (G043H7, Biolegend) CD14-V500 (M5E2; BD Bioscience), CD19-V500 (HIB19; BD Bioscience), CD25-FITC (M-A251; BD Bioscience), CD45RA-eFluor450 (HI100, eBiosciences), CD137-APC (4B4-1; BioLegend), OX40-PECy7 (Ber-ACT35; BioLegend), and PD-L1-PE (29E.2A3; BioLegend) for 20 min at room temperature. Cells were washed and resuspended in PBS before being transferred into a 5 ml polypropylene FACS tube (BD Bioscience). PBMCs were sorted, based on forward and side scatter excluding debris and doublets, on a FACSAria into TRIzol LS (Thermo Fisher). Acquisition files were analyzed using FlowJo. Total RNA was extracted from ∼100,000 cells in TRIzol LS using the miRNeasy Micro Kit (Qiagen) on a QIAcube (Qiagen). Total RNA was amplified according to Smart Seq protocol (2). cDNA was purified using AMPure XP beads. cDNA was used to prepare a standard barcoded sequencing library (Illumina). Samples were sequenced using an Illumina HiSeq2500 to obtain 50-bp single end reads. Samples that failed to be sequenced due to limited sample availability or failed the quality control were eliminated from further sequencing and analysis. The full protocol can be found at protocols.io (http://dx.doi.org/10.17504/protocols.io.bxr6pm9e).

### RNA-seq analysis

Paired-end reads that passed Illumina filters were filtered for reads aligning to tRNA, rRNA, adapter sequences, and spike-in controls. The reads were aligned to the GRCh38 reference genome and Gencode v27 annotations using STAR (v2.6.1) ^17^. DUST scores were calculated with PRINSEQ Lite v0.20.3 ^18^ and low-complexity reads (DUST > 4) were removed from BAM files. The alignment results were parsed via SAMtools ^19^ to generate SAM files. Read counts to each genomic feature were obtained with the featureCounts (v 1.6.5) ^20^ using the default option along with a minimum quality cut off (Phred > 10). After removing absent features (zero counts in all samples), the raw counts were imported into R v3.6.1 and genes with an average TPM < 1 were removed. R/Bioconductor package DESeq2 v.1.24.0 ^21^ was used to normalize raw counts. Variance stabilizing transformation was applied to normalized counts to obtain log_2_ gene expression values. Quality control was performed using boxplots and Principal component analysis (PCA), using the ‘prcomp’ function in R, on log_2_ expression values. Differentially expressed genes were identified using the DESeq2 Wald test, and p-values were adjusted for multiple test correction using the Benjamini Hochberg algorithm ^22^. Genes with adjusted p values < 0.05 and log2 fold change >1 or < -1 were considered differentially expressed. Cell type enrichment was performed using DICE ^23^. The RNAseq data have been submitted to the Gene Expression Omnibus under accession number GSE207214 (http://www.ncbi.nlm.nih.gov/geo/).

## Results

### HLA DQA1*03:01, DRB4*01:01, and DPB1*04:02 are associated with susceptibility for active TB and DPB1*105:01 with protection

To determine whether specific HLA alleles are associated with active TB, samples were obtained from a cohort of 636 participants, of which 203 individuals were QFT negative, the remaining 433 individuals were QFT positive including 99 individuals with a history of active TB (ATB) or who were diagnosed with active TB upon enrollment. Participants were HLA typed by determining the specific alleles expressed at the HLA-A, -B, -C, -DPA1, -DPB1, -DQA1, -DQB1,-DRB1, and -DRB3/4/5 loci. The results of the HLA typing were utilized to investigate whether the expression of particular HLA alleles would be associated with *Mtb* infection outcomes. To identify allele associations with TB, odds ratios and p-values between active TB and QFT negative/QFT positive individuals were calculated utilizing the RATE algorithm ^24^. A type 1 error rate of 0.05 was accounted for via a Bonferroni correction on alleles expressed in at least 5% of the total study cohort (**Table 1**).

**Table 1.** HLA allele association with active TB.

|  |  | No. participants<br>with allele |  | No. participants<br>lacking allele |  |  |  |  |  |
| --- | --- | --- | --- | --- | --- | --- | --- | --- | --- |
| HLA allele | HLA allele<br>frequency<br>(%) | Active<br>TB | QFT+<br>&<br>QFT- | Active<br>TB | QFT+<br>&<br>QFT- | Odds<br>ratio | P-value | Bonferroni<br>corrected<br>p-value | Population<br>coverage<br>(%) |
| DQA1*03:01 | 7.1 | 24 | 21 | 75 | 516 | 7.9 | $8.0\text{E-}10$ | **6.9E-08** | 26.4 |
| DPB1*04:02 | 10.3 | 25 | 40 | 74 | 495 | 4.2 | $2.0\text{E-}06$ | **1.7E-04** | 19.2 |
| DRB4*01:01 | 27.0 | 42 | 127 | 57 | 401 | 2.3 | 0.0003 | **0.03** | 47.4<sup>1</sup> |
| DPB1*105:01 | 25.6 | 12 | 150 | 87 | 385 | 0.4 | 0.0006 | **0.05** | 0.1<sup>2</sup> |
| A*23:17 | 6.8 | 0 | 43 | 99 | 494 | 0.0 | 0.0008 | >0.05 | 0.0<sup>3</sup> |
| DQA1*01:05 | 5.3 | 0 | 34 | 99 | 503 | 0.0 | 0.006 | >0.05 | 2.3 |
| DRB1*11:01 | 12.9 | 21 | 61 | 78 | 474 | 2.1 | 0.01 | >0.05 | 10.5 |
| DQA1*03:03 | 21.7 | 13 | 125 | 86 | 412 | 0.5 | 0.02 | >0.05 | 6.9 |
| B*15:03 | 9.9 | 16 | 47 | 83 | 489 | 2.0 | 0.03 | >0.05 | 1.3 |
| DQB1*02:01 | 15.4 | 8 | 90 | 90 | 447 | 0.4 | 0.03 | >0.05 | 23.0 |
| DRB1*03:01 | 17.5 | 10 | 101 | 89 | 434 | 0.5 | 0.04 | >0.05 | 17.8 |
| DRB1*07:01 | 17.8 | 25 | 88 | 74 | 447 | 1.7 | 0.04 | >0.05 | 18.2 |

<sup>1</sup> Coverage based in linkage with DR4, 7, and 9 alleles. <sup>2</sup> Rare worldwide, but 25.4% in South African Black and 20% in Brazilian Black. <sup>3</sup> Rare worldwide, but 7.7% in South African Black and 8% in Brazilian Black.

Of the 324 HLA alleles expressed in one or more individuals of our cohort, we focused on the 87 alleles (37 class I and 50 class II alleles) that were present in at least 5% (i.e. 32 individuals) of the cohort to ensure we had sufficient power to detect differences in the HLA distribution. For these 87 alleles, 12 had a significant p-value when comparing their frequency in individuals with ATB compared to QFT negative and positive individuals (**Table 1**). Most of these variants are commonly expressed in the general population (Table 1). Of these variants, HLA DQA1*03:01 had the strongest association with ATB, with an odds ratio of 7.9 and p-value of 8.0E-10. A total of 45 participants carried this allele type, 24 of which (53%) had ATB. In addition, the DPB1*04:02 and DRB4*01:01 alleles were also associated with ATB, with odds ratios of 4.2 and 2.3 respectively. Finally, HLA DPB1*105:01 appeared to confer protection against ATB, since it was inversely correlated with ATB (OR= 0.4, p=0.0006). After Bonferroni correction, both the positive association between alleles DQA1*03:01, DPB1*04:02, DRB4*01:01 and ATB (p= 6.9E-8, 1.7E-4, 0.03), and the negative association between HLA DPB1*105:01 and ATB (p=0.05) were maintained.

### DQA1*04:01 and DRB1*03:02 are associated with protection from *Mtb* infection

To protect from active disease, a factor like HLA could either protect from infection, or from progression to active TB. To investigate that we examined the alleles that were associated with susceptibility to active disease, and tested if these were enriched within the QFT positive. We found no alleles that were more frequent in QFT positive individuals compared to QFT negative (not shown).

We next investigated the differences in HLA expression between *Mtb* infected (both QFT positive and ATB) participants and QFT negative participants, using the same approach as above (**Table 2**). We identified 19 alleles present in at least 5% of the participants with a significant p-value. In this comparison, DQA1*03:01 was again identified to have the closest association to *Mtb* infection with an odds ratio of 11.1 and a corrected p-value of 0.001. In addition, DQA1*04:01 and DRB1*03:02 were associated with protection from *Mtb* infection, both with odds ratios of 0.4 (p=0.033 and 0.050, respectively). In conclusion, we identified several alleles associated with ATB and *Mtb* infection susceptibility. We focused the rest of the study on the alleles associated with susceptibility for ATB.

**Table 2.** HLA allele association with *Mtb* infection.

|  |  | No. participants<br>with allele |  | No. participants<br>lacking allele |  |  |  |  |
| --- | --- | --- | --- | --- | --- | --- | --- | --- |
| HLA allele | HLA allele<br>frequency<br>(%) | Active<br>TB &<br>QFT+ | QFT- | Active<br>TB &<br>QFT+ | QFT- | Odds<br>ratio | P-value | Bonferroni<br>corrected<br>p-value |
| DQA1*03:01 | 7.1 | 43 | 2 | 390 | 201 | 11.1 | 1.00E-05 | 0.001 |
| DQA1*04:01 | 13.1 | 42 | 41 | 391 | 162 | 0.4 | 3.80E- | 0.033 |
|  |  |  |  |  |  |  | 04 |  |
| **DRB1\*03:02** | 10.9 | 34 | 35 | 398 | 167 | 0.4 | 0.001 | **0.050** |
| DQB1*04:02 | 14.8 | 50 | 44 | 382 | 159 | 0.5 | 0.001 | >0.05 |
| DQA1*01:02 | 45.9 | 217 | 75 | 216 | 128 | 1.7 | 0.002 | >0.05 |
| C*17:01 | 14.2 | 49 | 41 | 384 | 161 | 0.5 | 0.003 | >0.05 |
| A*74:01 | 5.8 | 33 | 4 | 400 | 199 | 4.1 | 0.003 | >0.05 |
| B*42:01 | 8.8 | 28 | 28 | 404 | 175 | 0.4 | 0.004 | >0.05 |
| DRB1*15:03 | 18.0 | 90 | 24 | 342 | 178 | 2.0 | 0.006 | >0.05 |
| DQB1*06:02 | 37.3 | 177 | 60 | 255 | 143 | 1.7 | 0.006 | >0.05 |
| DPA1*02:02 | 37.1 | 135 | 61 | 191 | 142 | 1.6 | 0.010 | >0.05 |
| DRB4*01:01 | 27.0 | 128 | 41 | 298 | 160 | 1.7 | 0.012 | >0.05 |
| A*30:01 | 12.1 | 43 | 34 | 390 | 169 | 0.5 | 0.018 | >0.05 |
| DQA1*01:03 | 20.6 | 78 | 53 | 355 | 150 | 0.6 | 0.021 | >0.05 |
| B*15:03 | 9.9 | 51 | 12 | 381 | 191 | 2.1 | 0.022 | >0.05 |
| DRB3*01:01 | 28.7 | 110 | 70 | 316 | 131 | 0.7 | 0.023 | >0.05 |
| DPB1*04:01 | 24.9 | 96 | 62 | 335 | 141 | 0.7 | 0.030 | >0.05 |
| DRB1*07:01 | 17.8 | 67 | 46 | 365 | 156 | 0.6 | 0.034 | >0.05 |
| DRB1*11:01 | 12.9 | 64 | 18 | 368 | 184 | 1.8 | 0.042 | >0.05 |

### DQB1*02:02, *03:03, *04:02, *06:01/02/09 co-expression with DQA1*03:01 increases susceptibility to ATB

Mature HLA DQ molecules expressed on the cell surface are a dimer of DQ alpha and beta chains, and particular DQA alleles are preferentially co-expressed with particular DQB allelic variants. For this reason, we next sought to determine which DQ beta chain variants are most likely to be co-expressed with HLA DQA1*03:01 in ATB cases. To do so, we calculated the odds ratios and p-values of HLA DQ alpha and beta pairs to identify which pair is more closely associated to ATB (**Table 3**). Using the most prevalent DQ beta chains in our cohort, we paired various beta chains with DQA1*03:01 and determined the odds ratios with a confidence interval of 95%. When paired with beta chains DQB1*02:02, *04:02, *03:03, and *06:01/02, the odds ratios significantly increased from 7.9 to 54.2 (DQB1*02:02), 35.0 (DQB1*04:02), 8.4 (DQB1*03:03), 9.5 (DQB1*06:01), and infinity (DQB1*06:02; e.g. no expression in QFT positive or negative individuals), respectively. The odds ratio of association when DQA1*03:01 is paired to all possible DQB1 chains combined is 16, an increase from the original 7.9. Thus, when DQA1*03:01 is co-expressed with the beta chains highlighted in table 2, the association of DQA1*03:01 with ATB becomes even more pronounced.

**Table 3.** DQB1 alleles expressed with DQA1*03:01.

| DQA1 | DQB1 | No. ATB<br>w allele | No. ATB<br>w/o<br>allele | No.<br>QFT+QFT-<br>w allele | No.<br>QFT+QFT-<br>w/o allele | OR | p-value |
| --- | --- | --- | --- | --- | --- | --- | --- |
| DQA1*03:01 | - | 24 | 75 | 21 | 516 | 7.9 | 8.0E-10 |
| DQA1*03:01 | DQB1*02:02 | 9 | 89 | 1 | 536 | 54.2 | 3.1E-07 |
| - | DQB1*02:02 | 23 | 75 | 85 | 452 | 1.6 | 0.08 |
| DQA1*03:01 | DQB1*04:02 | 6 | 92 | 1 | 536 | 35.0 | 7.2E-05 |
| - | DQB1*04:02 | 11 | 87 | 83 | 454 | 0.7 | 0.35 |
| DQA1*03:01 | DQB1*06:02 | 8 | 90 | 5 | 532 | 9.5 | 1.6E-04 |
| - | DQB1*06:02 | 44 | 54 | 193 | 344 | 1.5 | 0.11 |
| DQA1*03:01 | DQB1*03:03 | 3 | 95 | 2 | 535 | 8.4 | 0.03 |
| - | DQB1*03:03 | 5 | 93 | 23 | 514 | 1.2 | 0.79 |
| DQA1*03:01 | DQB1*06:01 | 2 | 96 | 0 | 537 | inf | 0.02 |
| - | DQB1*06:01 | 8 | 90 | 32 | 505 | 1.4 | 0.37 |
| DQA1*03:01 | DQB1*06:09 | 1 | 97 | 0 | 537 | inf | 0.15 |
| - | DQB1*06:09 | 6 | 92 | 47 | 490 | 0.7 | 0.55 |
| DQA1*03:01 | DQB1<br>Combined | 21 | 77 | 9 | 528 | 16.0 | 0.26 |

### Expression of susceptibility alleles is associated with lower magnitude of *Mtb*-specific response in an IFNγ ELISPOT assay

We next investigated if the expression of the HLA alleles associated with ATB (**Table 1**) was associated with differences in the *Mtb*-specific immune response. PBMC samples from a subset of our cohort (all of which were QFT positive and 59 of which were ATB cases) were tested for IFNγ production in response to an *Mtb*-derived peptide pool (MTB300). For all three of the susceptibility alleles, (DQA1*03:01, DPB1*04:02, or DRB4*01:01) we found a trend towards a lower magnitude of IFNγ-specific responses, with the strongest trend being for DQA1*03:01 (**Figure 1a-c**). When the three alleles were considered together, we found a significantly lower magnitude of IFNγ-specific response in individuals with the susceptibility alleles compared to those without (**Figure 1d**). No significant difference was detected when comparing the responses from the ATB cohort with the QFT positive cohort (not shown). In conclusion, the expression of the three susceptibility alleles influences the *Mtb*-specific IFNγ response.

**Figure 1.**
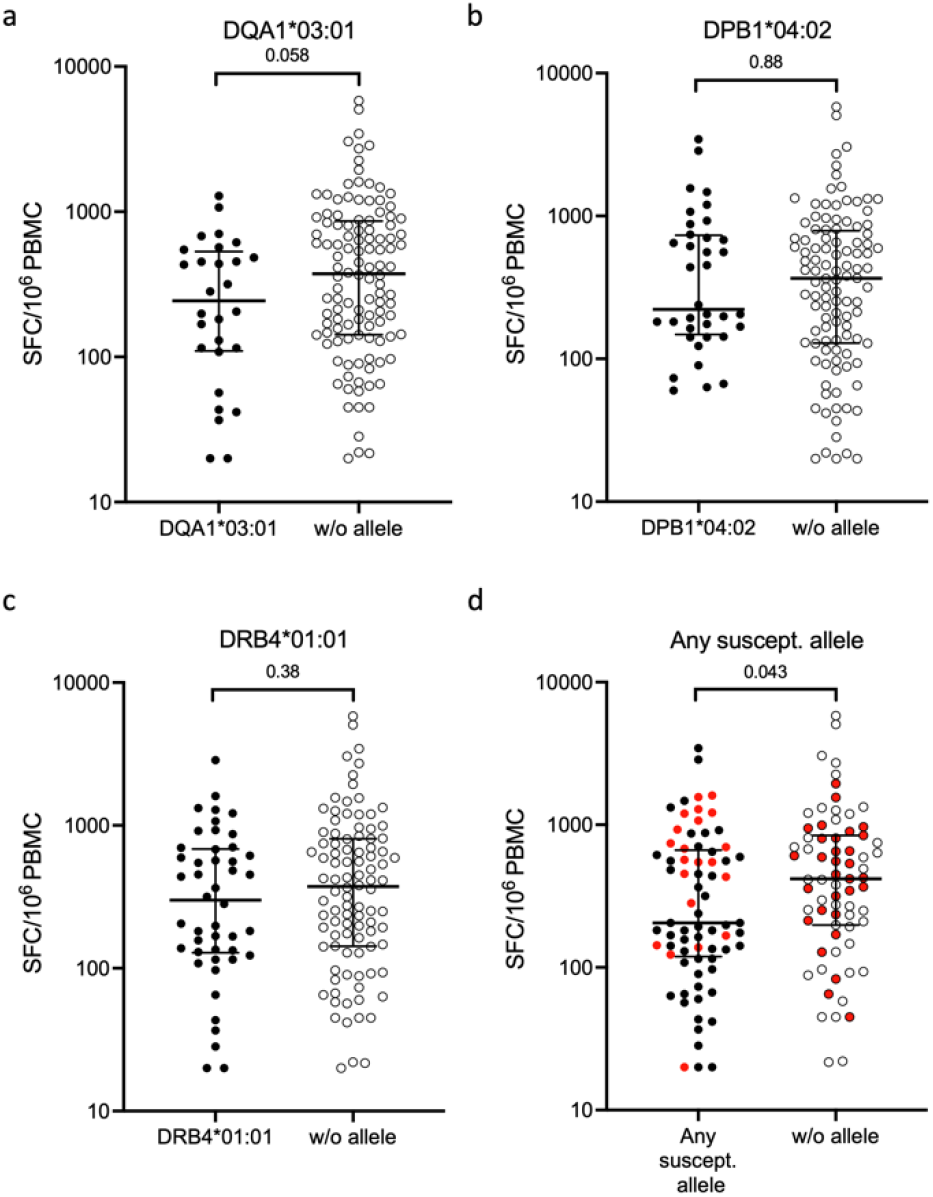
The expression of susceptibility alleles is associated with a lower magnitude of *Mtb*-specific IFNγ response. Magnitude of response against MTB300 as SFC per 10^6^ PBMC measured in an IFNγ ELISPOT assay. Each point represents one individual (total n=144), median ± interquartile range is shown. Two-tailed Mann-Whitney U test. Individuals with susceptibility alleles are represented by closed circles and individuals without by open circles. a) DQA1*03:01 n=28 with and n=116 without, b) DPB1*04:02 n=36 with, n=108 without, c) DRB4*01:01 n=46 with, n=98 without, d) any susceptibility allele n=73 with, n=71 without. Red color indicates participants included in subsequent gene expression experiments.

### The presence of ATB susceptibility alleles is correlated with specific gene signatures

To further investigate the biological mechanisms of HLA association with ATB susceptibility, PBMCs from a subset of QFT positive healthy individuals (**Fig 1d**, red color) were stimulated with MTB300 or a whole cell lysate (WCL) of the H37Rv TB strain. Live, singlet cells were sorted and their global gene expression was determined by RNA-Seq. The differential gene expression analysis of individuals who express the susceptibility alleles vs. individuals who lack the expression of these alleles in each stimulation condition from the ELISPOT assay (unstimulated, MTB300, and WC), revealed a specific gene signature.

A total of 84 differentially expressed genes (DEGs) were identified across the three different stimulation conditions (**Supplementary table 2**; adjusted p values <0.05 and log2 fold change >1 or <-1). For the unstimulated samples, 5 genes were upregulated in individuals with the susceptibility alleles, and 29 genes were upregulated in those without. For the MTB300 stimulation condition, 8 genes were upregulated in with and 10 w/o, and for WCL it was 21 vs. 19 genes (**Figure 2a**). The 84 genes were filtered based on concordant expression across the different stimuli, e.g. if a gene was upregulated in individuals with the susceptibility allele in one condition but up in individuals without in another it was removed. This resulted in a gene signature of 63 genes (**Figure 2b and Supplementary table 2**). These genes could be grouped into 4 clusters based on their expression in individuals with vs. without the susceptibility alleles and across stimuli conditions (**Figure 2b**). Cluster 1 encompasses genes that were upregulated in the unstimulated and MTB300 condition, but with no difference between allele expression. The genes corresponding to Cluster 2 were upregulated in individuals with expression of the susceptibility alleles across the three different conditions compared to individuals lacking the alleles. Cluster 3 consisted of genes upregulated in individuals without the susceptibility alleles across different stimulations. Interestingly, cluster 3 included HLA-DQA1 and HLA-DQB1, as well as JUN and TIRAP. Cluster 4 was composed of genes with different patterns of upregulation in response to WCL, but no difference between allele expression.

**Figure 2.**
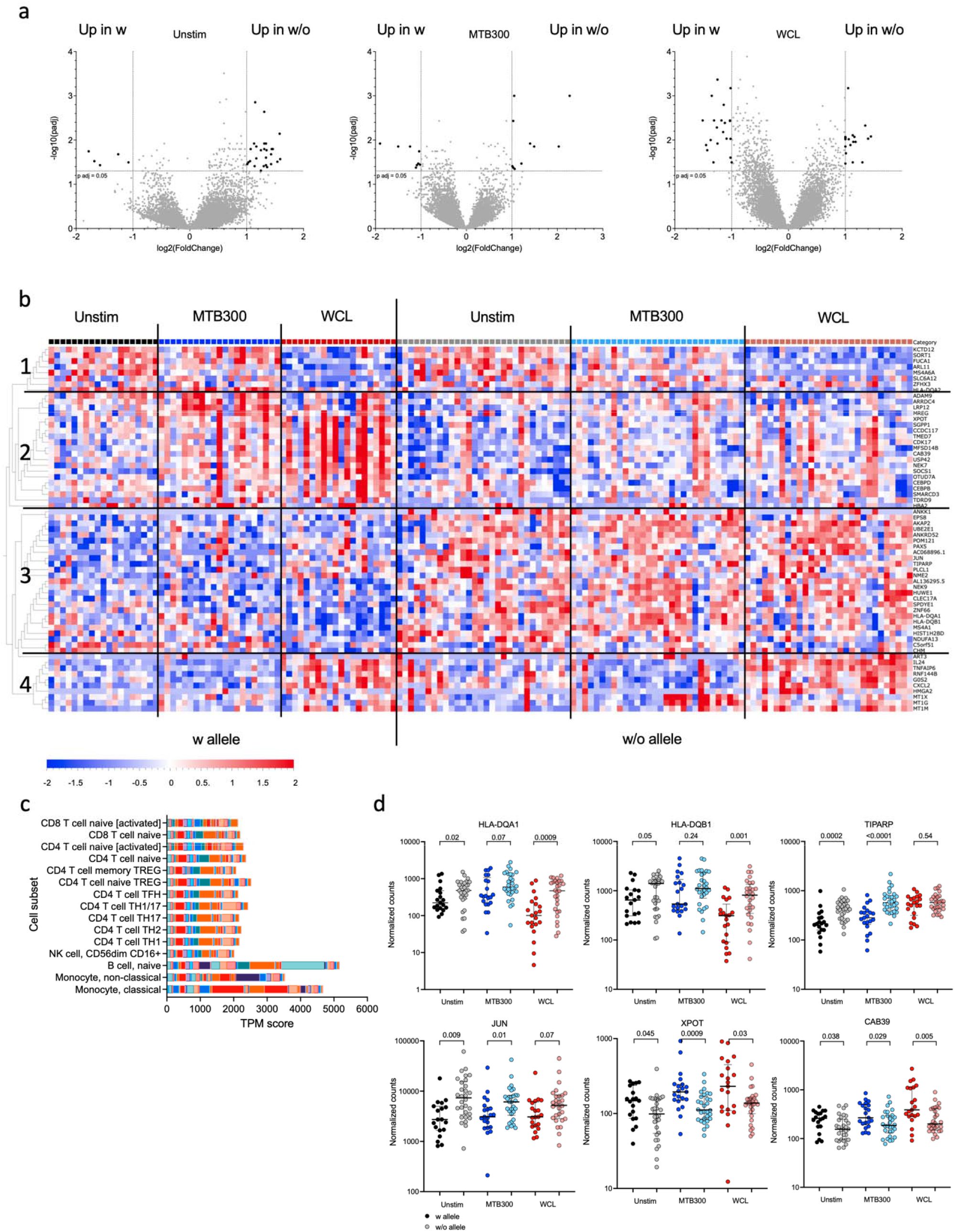
Expression of the susceptibility alleles is associated with a specific gene signature. a) Volcano plots showing differentially expressed genes comparing individuals with susceptibility alleles (n=21) to those without (n=30) in three different stimuli conditions; unstimulated, MTB300, and WCL. Black indicates DEGs (adjusted p value <0.05 and log2 fold change >1 and <-1). b) Heatmap displaying the 63 DEGs from (a). Genes are ordered by hierarchical clustering, and samples are ordered according to allele expression and then according to stimulation condition. Black vertical lines indicate allele and stimuli separation and horizontal lines indicate cluster separation. c) Cell-type enrichment (dice-database.org) for the 63 DEGs. Each color represents one gene. d) Gene expression at the mRNA level. Gene expression values in counts normalized by sequencing depth calculated by the DEseq2 package. Two-tailed Mann-Whitney U test.

The 63 genes associated with the signature were enriched in genes expressed in B cells and monocytes (both classical and non-classical; **Figure 2c**). Finally, to visualize the gene expression changes between cohorts in each stimulus, 6 representative genes were chosen, 2 from cluster 2 (XPOT and CAB39) and 4 from cluster 3 (JUN, TIPARP, HLA-DQA1, HLA-DQB1; **Figure 2d**). The 4 genes from cluster 3 exhibited a trend for lower expression in individuals with the susceptibility alleles in each of the stimuli conditions. Conversely, the 2 genes from cluster 2 had higher expression in each stimuli condition in individuals with the susceptibility alleles.

### Gene signatures associated with ATB susceptibility alleles and low T cell responsiveness to *Mtb* antigens

Finally, we investigated whether DEGs could be identified in PBMCs from participants who had different magnitudes of an *Mtb*-specific IFNγ response. Specifically, we compared the unstimulated samples from individuals expressing any of the three susceptibility alleles and who had a magnitude of response below the median for individuals included in the RNAseq (<568 SFCs) with individuals lacking those alleles and who had a magnitude of response above the median. Unstimulated samples were used for this analysis to avoid detecting genes differentially expressed solely due to the *Mtb*-specific magnitude i.e. potentially higher levels of T cell activation genes, etc. This analysis revealed 21 DEGs which were all upregulated in individuals without the susceptibility alleles (**Figure 3a**). This gene signature could distinguish between these two cohorts (with alleles; low expression and without alleles; high expression, **Figure 3b**,**c**). Furthermore, 5 of these 21 DEGs overlapped with the 63-gene signature described above (CXCL2, JUN, TNFAIP6, ZNF66, and AC068896.1).

**Figure 3.**
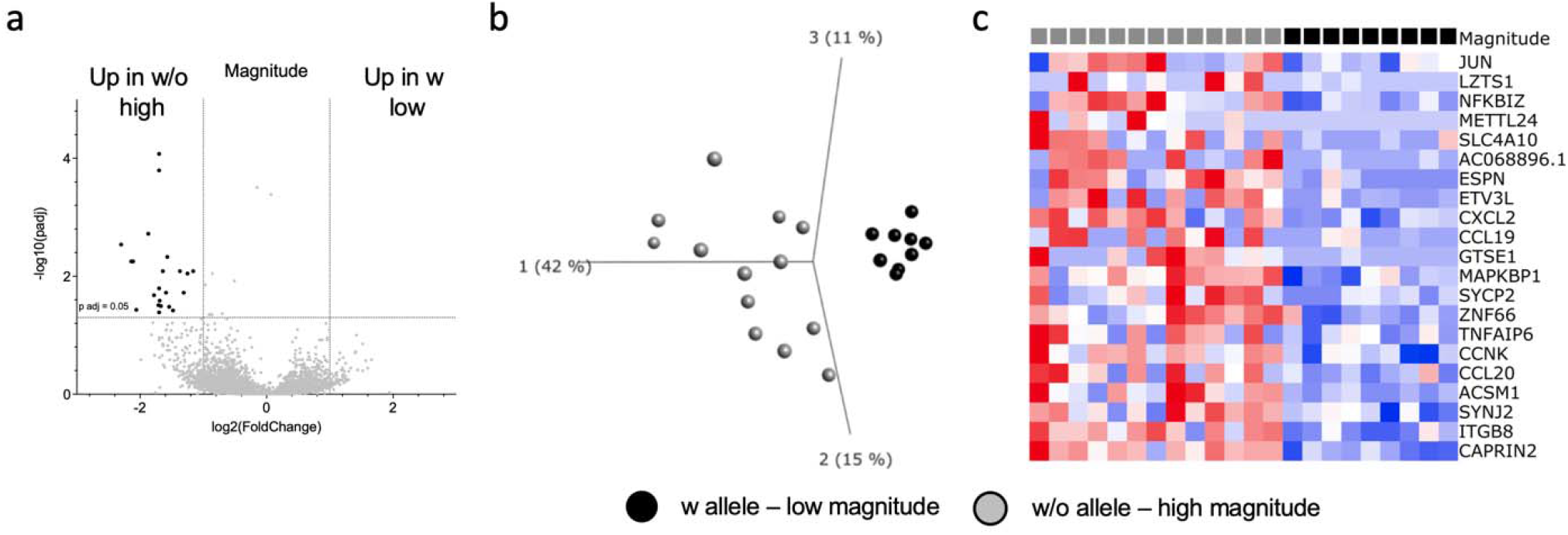
Differences in *Mtb*-specific magnitude of response is associated with a specific gene signature. a) Volcano plot showing differentially expressed genes comparing individuals with susceptibility alleles and low magnitude of expression (n=9) to those without and high magnitude of expression (n=13) in unstimulated samples. Black indicates DEGs (adjusted p value <0.05 and log2 fold change >1 and <-1). b) PCA plot illustrating differences between the two cohorts for the 21 DEGs. c) Heatmap displaying the 20 DEGs from (a). Genes are ordered by hierarchical clustering, and samples are ordered according to the two cohorts.

Taken together, individuals expressing any of the susceptibility alleles have a specific gene signature that is enriched in genes related to APCs, possibly related to the lower magnitude of *Mtb*-specific IFNγ responses detected.

## Discussion

In this study, we examined the potential association between the expression of specific HLA alleles and ATB disease. The identification of risk factors for ATB will help identify individuals at high risk of progression and could inform preventative treatment strategies. Using samples from 636 participants, we identified 3 HLA alleles associated with susceptibility for ATB. The expression of these alleles was associated with a lower magnitude of *Mtb*-specific T cell responses and also correlated with a lower expression of APC related genes.

HLA alleles are highly polymorphic in general, and the genetic heterogeneity in HLA alleles also varies across different ethnic groups and geographical populations. However, the alleles identified as associated with differential disease outcomes in the present study are relatively common in most ethnic groups. While we identified an association of ATB with DQA1*03:01 in our study cohort from South Africa and the US, this allele was also found in association with pulmonary tuberculosis in a cohort from Iceland and Russia ^25^. We further identified the DQB1 alleles most likely expressed together with DQA1*03:01. These DQB1 alleles were DQB1*02:02, *04:02, *03:03, and *06:01/02. A meta-analysis of 12 studies found that HLA DQB1*06:01 was significantly associated with an increased risk of ATB as well ^26^.

Sveinbjornsson et al. reported the association of the HLA class II allele DQA1*03 with *Mtb* infection and identified a noncoding variant, rs557011, located between the HLA-DQA1 and HLA-DRB1 alleles that also associated with *Mtb* infection ^25^. Many other groups have reported similar findings for different class I and class II HLA alleles ^27-30^. Due to the polymorphic nature and heterogeneity of HLA allele expression in diverse geographical locations, further studies in additional cohorts across different ethnicities may identify or confirm additional HLA alleles associated with susceptibility to ATB. In addition, HLA alleles present a strong linkage disequilibrium in which preferential combinations of alleles are inherited together in the genome more often than expected. The series of alleles at linked loci on a single chromosome is named “haplotype” ^31^. Specific haplotypes of association with ATB may be identified through more detailed HLA typing in which these linked loci are taken into consideration. In our study we did not determine these linkages.

We further linked expression of the susceptibility alleles to changes in the *Mtb*-specific immune response. Individuals expressing DQA1*03:01, DPB1*04:02, and DRB4*01:01 had a lower *Mtb*-specific immune response compared to those lacking these alleles. Other groups have reported impaired immune responses due to the expression of specific HLA alleles in patients with human immunodeficiency virus (HIV) and hepatitis c virus (HCV) infection ^32-35^. Huang et al. identified an HLA-B*35 variant that was associated with accelerated HIV infection in a population of patients of European descent. They described that individuals who expressed the HLA-B*35 susceptibility allele had significantly impaired dendritic cell (DC) function than individuals lacking the susceptibility allele and hypothesized that this impaired DC function was the mechanism by which the susceptibility allele contributed to HIV disease ^33^. Additionally, HLA class I and class II variants have been found to confer protection against severe HCV infection and these protective alleles have been reported to better present immunodominant HCV epitopes ^32,34,35^. Finally, HLA-DQ variants have been reported to present streptococcal and staphylococcal superantigens at different strengths and influence the overall T cell-mediated response to bacterial superantigens ^36^. Overall, HLA allele variants have been linked previously to altered immune responses in the context of different diseases.

In addition, using transcriptomics we defined gene signatures that could distinguish individuals with vs. without expression of the susceptibility alleles. The DEGs could be divided into 4 clusters that provided further insight into the role of the HLA susceptibility alleles and *Mtb* infection. Two clusters corresponded to genes that were differentially expressed in response to the specific stimuli unstim and MTB300 vs. WCL stimulation. One cluster was comprised of genes that were upregulated in individuals who express the susceptibility alleles. These genes included the transcription factors CEBPD and CEBPB, which are highly expressed on monocytes (www.dice.org and www.proteinatlas.org). Finally, the last gene cluster was comprised of genes that were upregulated in individuals who are not carriers of the susceptibility alleles. This cluster included both HLA-DQA1 and HLA-DQB1, as well as PAX5, JUN and CLEC17A which are all highly expressed on B cells (www.dice.org). JUN is involved in toll-like receptor activity and antigen-cross presentation ^37^, and downregulation (as in individuals expressing the susceptibility alleles) leads to impairment of those functions. This cluster also contained TIRAP, which encodes for a molecule that is associated with toll-like receptors and also plays an important role in innate immunity ^38^. Thus, individuals expressing the susceptibility alleles have downregulation of important immune response related genes, which may contribute to an increased risk of ATB.

Lastly, we identified a gene signature related to the magnitude of *Mtb*-specific T cell responses. This signature was upregulated in individuals lacking the susceptibility alleles and 5 genes overlapped with the signature detected when comparing DEGs irrespective of the *Mtb*-specific T cell response detected. This signature included JUN, as mentioned above, as well as, CCL19 which is involved in the homing of lymphocytes to the lung in *Mtb* infection ^39^. The signature also included CXCL2 which is a neutrophil-attracting chemokine ^40^. Taken together, these gene signature results suggest that the susceptibility alleles potentially increase the risk for ATB by impairing antigen loading and presentation to immune cells for proper clearance of *Mtb* infection.

In conclusion, our study identified three HLA alleles (HLA DQA1*03:01, DPB1*04:02, and DRB4*01:01) associated with susceptibility to ATB. Further investigation revealed that individuals expressing these alleles have a decreased *Mtb*-specific immune response. The identified HLA alleles and associated gene signatures can help identify individuals who are at risk of ATB.

## Data Availability

All data produced in the present study are available upon reasonable request to the authors.

## Author contributions

TJS, AS, BP, and CSLA participated in the design and direction of the study. LYC, RK, and CSLA performed literature search and participated in data collection, analysis and interpretation. VR and TJS and the SATVI-Ja Jolla Study Group recruited participants. EJP, SAM, and MMD coordinated and performed HLA typing. LYC and CSLA wrote the first draft. All authors reviewed, edited and approved the final version of the manuscript before submission.

## Declaration of interests

The authors declare no competing interests.

## Acknowledgements

We thank the sequencing core and flow cytometry core at La Jolla Institute for Immunology for their help in these studies.

This study was supported by the National Institutes of Health grants U19 AI118626, S10 RR027366, S10 OD016262.

## The SATVI study group

|  |  |
| --- | --- |
| Thomas Scriba | PI |
| Michelle Fisher | Project Manager |
| Humphrey Mulenga | Data Manager |
| Ashley Veldsman | Regulatory Manager |
| Nicole Bilek | Laboratory Manager |
| Angelique Luabeya | Site PI |
| Jateel Kassim | Site PI (Previous) |
| Marwou de Kock | Site Manager |
| Cynthia Ontong | Study Co-ordinator |
| Elizabeth Filander | Research Nurse |
| Gail Jacobs | CRW |
| Nambitha Nqakala | CRW |
| Yulandi Herselman | Data Administrator |
| Simba Mabwe | Site Lab Manager |
| Denis Arendsen | Lab Technologist (study supervisor) |
| Palesa Dolo | Lab Technologist |
| Hadn Africa | Lab Technologist (Expert knowledge regarding processing with blood bags) |
| Marcia Steyn | Lab Technologist |
| Lebohang Makhethe | Lab Technologist |

## Notes

### Competing Interest Statement

The authors have declared no competing interest.

